# SARS-CoV-2 shedding and evolution in immunocompromised hosts during the Omicron period: a multicenter prospective analysis

**DOI:** 10.1101/2023.08.22.23294416

**Authors:** Zoe Raglow, Diya Surie, James D. Chappell, Yuwei Zhu, Emily T. Martin, Jennie H. Kwon, Anne E. Frosch, Amira Mohamed, Julie Gilbert, Emily E. Bendall, Auden Bahr, Natasha Halasa, H. Keipp Talbot, Carlos G. Grijalva, Adrienne Baughman, Kelsey N. Womack, Cassandra Johnson, Sydney A. Swan, Emilia Koumans, Meredith L. McMorrow, Jennifer L. Harcourt, Lydia J. Atherton, Ashley Burroughs, Natalie J. Thornburg, Wesley H. Self, Adam S. Lauring

**Author notes:** For the Investigating Respiratory Viruses in the Acutely Ill (IVY) Network. **Corresponding Author:** Adam S. Lauring 1137 Catherine Street, MS2 4742C Ann Arbor, MI 48109.

## Abstract

**Background:** Prolonged SARS-CoV-2 infections in immunocompromised hosts may predict or source the emergence of highly mutated variants. The types of immunosuppression placing patients at highest risk for prolonged infection and associated intrahost viral evolution remain unclear.

**Methods:** Adults aged ≥18 years were enrolled at 5 hospitals and followed from 4/11/2022 – 2/1/2023. Eligible patients were SARS-CoV-2-positive in the previous 14 days and had a moderate or severely immunocompromising condition or treatment. Nasal specimens were tested by rRT-PCR every 2–4 weeks until negative in consecutive specimens. Positive specimens underwent viral culture and whole genome sequencing. A Cox proportional hazards model was used to assess factors associated with duration of infection.

**Results:** We enrolled 150 patients with: B cell malignancy or anti-B cell therapy (n=18), solid organ or hematopoietic stem cell transplant (SOT/HSCT) (n=59), AIDS (n=5), non-B cell malignancy (n=23), and autoimmune/autoinflammatory conditions (n=45). Thirty-eight (25%) were rRT-PCR-positive and 12 (8%) were culture-positive ≥21 days after initial SARS-CoV-2 detection or illness onset. Patients with B cell dysfunction had longer duration of rRT-PCR- positivity compared to those with autoimmune/autoinflammatory conditions (aHR 0.32, 95% CI 0.15-0.64). Consensus (>50% frequency) spike mutations were identified in 5 individuals who were rRT-PCR-positive >56 days; 61% were in the receptor-binding domain (RBD). Mutations shared by multiple individuals were rare (<5%) in global circulation.

**Conclusions:** In this cohort, prolonged replication-competent Omicron SARS-CoV-2 infections were uncommon. Within-host evolutionary rates were similar across patients, but individuals with infections lasting >56 days accumulated spike mutations, which were distinct from those seen globally.

## INTRODUCTION

Over the past three years, the COVID-19 pandemic has been characterized by the emergence of highly mutated variants of concern (VOC) with altered transmissibility, virulence, and/or ability to evade neutralization by therapeutic or vaccine-induced antibodies [1,2].

Immunocompromised patients are central to many of the clinical and epidemiologic aspects of SARS-CoV-2 VOC; they are less protected by vaccines [3,4] and may not develop sufficient immunity to clear the virus, even in the presence of monoclonal antibodies or antiviral drugs [5,6].

Early studies suggested that many immunocompromised individuals are at risk for prolonged infection with SARS-CoV-2 [7–9]. Among hospitalized patients, those with immunocompromising conditions are more likely to have detectable viral RNA and to be viral culture positive beyond 21 days [10–12]. Individuals with hematologic malignancy [10] and people living with AIDS [13,14] appear to be at greatest risk for prolonged infection. A large number of case reports of single [7–9,15–17] and multiple [18–20] patients have documented that a subset of immunocompromised patients are at risk for very prolonged infections, lasting hundreds of days. Because nearly all these studies are retrospective, with varying levels of ascertainment bias, prospective studies are needed to fully define this problem and those most at risk.

While the propagation of novel SARS-CoV-2 mutations is generally limited by host clearance and the stochastic dynamics of transmission [21–23], extended within-host replication in immunocompromised hosts allows the virus sufficient time to accumulate mutations. If transmitted [24], these viruses will appear to have evolved at an “accelerated rate” with more mutations per unit time [25]. The increasing identification of multi-mutational events in immunocompromised hosts and the abrupt emergence of highly mutated VOC have led to the hypothesis that the Alpha (B.1.1.7) and Omicron (BA.1) variants, and perhaps other VOCs, originated during these very prolonged infections within immunocompromised individuals [9,17,26–28]. This hypothesis is further supported by the selection of immune escape mutations in immunocompromised patients treated with convalescent plasma [9,26]. Importantly, many of the reported cases of extensive within-host evolution originated in the pre-Alpha or early Alpha variant era, prior to the introduction of vaccines and more effective antivirals. It is therefore unclear whether current interventions will limit—or, alternatively, drive—the evolution of highly mutated variants in these individuals and whether this pattern will be replayed on the Omicron genetic background.

To address ongoing and urgent questions related to SARS-CoV-2 infections in immunocompromised hosts, we performed prospective surveillance in immunocompromised inpatients and outpatients diagnosed with Omicron variant SARS-CoV-2 infection. Adult patients were enrolled at 5 sites in the Investigating Respiratory Viruses in the Acutely Ill (IVY) Network, a collaboration with the US Centers for Disease Control and Prevention (CDC) [4,29]. Through analysis of detailed clinical, RNA viral load, viral culture, and sequence data from prospectively collected specimens, we define those most at risk for prolonged infection among the cohort studied, the impact of antiviral treatments, and SARS-CoV-2 evolutionary dynamics in an immunocompromised population.

## METHODS

This program was determined to be public health surveillance with waiver of participant informed consent by CDC and institutional review boards at all participating institutions and was conducted in accordance with applicable CDC policy and federal law (45 C.F.R. part 46.102(l)(2), 21 C.F.R. part 56; 42 U.S.C. §241(d); 5 U.S.C. §552a; 44 U.S.C. §3501 et seq).

### Participants

Immunocompromised adults aged ≥18 years with SARS-CoV-2 infection were enrolled from inpatient and outpatient settings at 5 sites in the IVY Network during April 11 – October 1, 2022. Patients were eligible for the study if they had a positive real-time reverse transcription PCR (rRT-PCR) test for SARS-CoV-2 within the previous 14 days collected as part of routine clinical care and a moderately or severely immunocompromising condition (https://www.covid19treatmentguidelines.nih.gov/special-populations/immunocompromised/). Enrolled patients were followed until they cleared SARS-CoV-2, as evidenced by 2 consecutive negative rRT-PCR tests approximately 2 weeks apart.

Patients were categorized into the following 5 groups based on their underlying immunosuppression: (1) B cell dysfunction, defined as patients receiving B cell depletion or chimeric antigen receptor T cell (CAR-T) therapy expected to have current activity on the patient’s immune system, and/or those with B cell malignancy or myeloma; (2) solid organ or hematopoietic stem cell transplant (SOT/HSCT), defined as patients with a history of solid organ or hematopoietic stem cell transplant and on immunosuppressive therapy; (3) acquired immune deficiency syndrome (AIDS), defined as patients with HIV and CD4 <200 cells/mcL or an AIDS-defining illness in the preceding 12 months; (4) malignancy, defined as patients with non-B cell malignancy on cytotoxic, myelosuppressive, or immunomodulatory chemotherapy; and (5) autoimmune/autoinflammatory, defined as patients with conditions treated with immunosuppression and not meeting criteria for another category [30]. See Supplemental Table 1 for a list of qualifying immunosuppressive medications.

### Data Collection

Demographic and clinical data were collected from electronic medical record review and patient (or proxy) interview and included: age, sex, race, ethnicity, underlying medical conditions, symptoms, COVID-19 vaccination history (vaccine type and dates of each dose), and treatment history, which captured immunosuppressant medications and treatment for SARS-CoV-2, including outpatient monoclonal antibody therapy and other antiviral therapies. Day zero for each patient was defined as the earliest of three dates: symptom onset, first positive SARS-CoV-2 test for the current episode of infection (included to capture patients with positive tests prior to study enrollment), or the most recent positive SARS-CoV-2 test that made the patient eligible for the study. This definition was chosen in order to approximate the true onset date of infection as closely as possible, as not all patients were symptomatic, and some had prolonged infection prior to study enrollment.

### Specimen Collection

Nasal swab specimens were collected from each participant at the time of enrollment and every 2-4 weeks thereafter until viral RNA-negative by rRT-PCR for two consecutive specimens. Specimens were initially preserved in viral transport media at 2 – 8°C and shipped to a central laboratory (Vanderbilt University Medical Center, Nashville, Tennessee) where they were aliquoted, tested by rRT-PCR, and stored at -70°C [4,29,31]. One aliquot was sent to the University of Michigan (Ann Arbor, Michigan) for whole genome sequencing and another to the Centers for Disease Control and Prevention (Atlanta, Georgia) for viral culture.

### RNA Viral Load and Viral Culture

RNA was extracted from 200µl of specimen transport media using the MagMax Viral/Pathogen II Nucleic Acid Isolation Kit on a KingFisher instrument, eluted in 50µl water, and stored at -70˚C. Amplification of total and subgenomic (sg) transcripts for nucleocapsid (N) genes was performed using amplification conditions described previously [31,32].

Specimens were cultured on Vero E6 cells (NR-54970, BEI Resources) stably overexpressing the transmembrane protease, serine 2 (TMPRSS2) and angiotensin-converting enzyme 2 (ACE2) using a previously described method [33].

### Viral Genomic Sequencing

Sequencing libraries were prepared using the NEBNext ARTIC SARS-CoV-2 Library Prep Kit and ARTIC v5.3.2 primer sets [34]. After barcoding, pooled libraries were size selected by gel extraction and sequenced on an Illumina Nextseq 1000 (P1 flow cell 2×300bp reads). Sequencing reads were aligned to the Wuhan-Hu-1 reference using BWA-mem v0.7.15 [35]. Primers were trimmed and consensus sequences were generated using iVar v1.2.1 [36]. We used PANGO and Nextclade to annotate SARS-CoV-2 lineages and clades, respectively [37]. Intrahost single nucleotide variants (iSNV) were identified using iVar [36] with the following criteria: frequency 0.02-1, p-value <1×10^-5^, variant position coverage depth >100x, variant allele read depth ≥10, variant quality score >35, and genome completeness >95%. We also masked ambiguous and homoplastic sites [38]. Indels were identified at the consensus (>50% frequency) level only. Only iSNV present at frequencies >2% were included in subsequent analyses; this threshold was chosen in order to limit false-positive mutations due to sequencing errors and/or library preparation. In analyses of within-host evolution, any mutation present at ≥98% frequency in the patient’s first sample was excluded in order to limit the contribution from fixed Omicron-related mutations.

Within-host divergence was calculated as in [39]. The frequencies of each type of mutation (synonymous, non-synonymous, and stop/nonsense) in each SARS-CoV-2 gene for every patient were summed. Within-host evolution was evaluated by comparing the first and last positive collected sample for each patient. The sum of the frequencies was then normalized to the number of sites available for each type of mutation in order to obtain a per-site viral divergence. The per-site viral divergence was divided by the number of days between specimen collection and infection onset date (day zero).

### Neutralization Assays

The focus reduction neutralization test (FRNT) assay for measuring SARS-CoV-2 neutralizing antibodies was adapted from [40]. Confluent Vero E6-TMPRSS2-T2A-ACE2 cells (NR-54970, BEI Resources) were utilized to characterize initial and evolved SARS-CoV-2 viruses against six sera pools created based on anti-spike IgG levels (BAU/ml) (V-PLEX SARS-CoV-2 Panel 2 Kit, Meso Scale Diagnostics, LLC).

### Statistical Analysis

We summarized participant characteristics using proportions (frequencies), means (with standard deviations), and medians (with interquartile ranges). Comparisons of demographic characteristics and COVID-19 vaccination status were performed using Chi-square or Wilcoxon/Kruskal-Wallis tests when appropriate. The alpha level was not adjusted for multiple comparisons, except where indicated.

We compared the duration of rRT-PCR positivity (the number of days from day zero to last SARS-CoV-2 positive test date) among different immunocompromised groups using a Cox proportional hazards model with the autoimmune/autoinflammatory group as the referent (as this group had the lowest level of immunosuppression). Covariates included age, sex, race/ethnicity, prior COVID vaccination, and antiviral use at baseline (defined as receipt of any antiviral between 90 days prior and 7 days after enrollment). Right censoring was applied for individuals who continued to test positive by rRT-PCR at 90 days. We used the log-rank test to compare differences among immunocompromising groups with p<0.05 considered statistically significant.

All analyses were conducted using R v4.1.3 (Boston, Massachusetts).

### Data Availability

Raw sequencing reads are available on the NCBI short read archive under BioProject PRJNA896930 and consensus sequences are available on GISAID.

## RESULTS

### Participants

During April 11, 2022 - October 1, 2022, 156 patients began enrollment procedures; 6 were excluded due to not completing enrollment procedures (3), not meeting eligibility criteria (2), or patient withdrawal (1), resulting in 150 patients in the final analysis (Supplementary Figure 1). Patient follow-up continued until February 1, 2023. Among the 150 patients, 59 (39%) were male, and the median age was 60 years (Table 1). COVID-19 vaccination status at enrollment included 12 (8%) unvaccinated, 4 (3%) who had received one vaccine dose, and 134 (89%) who had received at least 2 vaccine doses. Immunocompromised category included 18 (12%) in the B cell dysfunction group, 59 (39%) in the SOT/HSCT group, 5 (3%) in the AIDS group, 23 (15%) in the malignancy group, and 45 (30%) in the autoimmune/autoinflammatory group (Table 1, Supplemental Table 2). One hundred thirty-five (90%) were symptomatic at the time of enrollment. Median time from illness onset or initial positive SARS-CoV2 test to study enrollment was 5 days (IQR 3-11). One hundred eleven (74%) patients received antiviral treatment between 90 days prior to and 7 days after enrollment, including remdesivir, molnupiravir, nirmatrelvir/ritonavir, convalescent plasma, or any monoclonal antibody; 6 (4%) received tixagevimab/cilgavimab prophylaxis, 27 (18%) received bebtelovimab, 1 (0.7%) received sotrovimab, and 4 (2%) received convalescent plasma (Supplemental Table 2).

**Table 1.** Characteristics at enrollment of immunocompromised patients with SARS-CoV-2 infection — IVY Network, 5 U.S. States*, April 11, 2022 – February 1, 2023.

| Characteristic | All patients<br>(N=150)<br>n (%) | B cell dysfunction<br>(N=18)<br>n (%) | SOT/HSCT<br>(N=59)<br>n (%) | AIDS<br>(N=5)<br>n (%) | Malignancy<br>(N=23)<br>n (%) | Autoimmune/<br>Autoinflammatory<br>(N=45)<br>n (%) |
| --- | --- | --- | --- | --- | --- | --- |
| Median age, yrs (IQR) | 60 (46–68) | 64 (50–71) | 60 (46–68) | 46 (45–57) | 65 (53–70) | 57 (46–65) |
| Sex |  |  |  |  |  |  |
| Female | 91 (61) | 11 (61) | 27 (46) | 3 (60) | 15 (65) | 35 (78) |
| Male | 59 (39) | 7 (39) | 32 (54) | 2 (40) | 8 (35) | 10 (22) |
| Race and ethnicity |  |  |  |  |  |  |
| White, non-Hispanic | 88 (59) | 8 (44) | 33 (56) | 1 (20) | 17 (74) | 29 (64) |
| Black, non-Hispanic | 39 (26) | 5 (28) | 20 (34) | 2 (40) | 2 (9) | 10 (22) |
| Hispanic | 11 (7) | 3 (17) | 3 (5) | 0 (0) | 1 (4) | 4 (9) |
| Other race, Unknown | 12 (8) | 2 (11) | 3 (5) | 2 (40) | 3 (13) | 2 (4) |
| Presence of symptoms at enrollment |  |  |  |  |  |  |
| Asymptomatic | 15 (10) | 1 (6) | 8 (14) | 1 (20) | 2 (9) | 3 (7) |
| Symptomatic | 135 (90) | 17 (94) | 51 (86) | 4 (80) | 21 (91) | 42 (93) |
| Days from illness onset or positive SARS-CoV-2 test result to enrollment |  |  |  |  |  |  |
| Median days (IQR) | 5 (3–11) | 5 (3–14) | 5 (3–11) | 2 (2–3) | 4 (3–6) | 5 (3–9) |
| Number of COVID-19 vaccine doses received |  |  |  |  |  |  |
| 0 | 12 (8) | 2 (11) | 2 (3) | 1 (20) | 3 (13) | 4 (9) |
| 1 | 4 (3) | 0 (0) | 3 (5) | 0 (0) | 0 (0) | 1 (2) |
| 2 | 26 (17) | 1 (6) | 13 (22) | 1 (20) | 2 (9) | 9 (20) |
| 3 | 63 (42) | 11 (61) | 17 (29) | 1 (20) | 15 (65) | 19 (42) |
| 4 | 37 (25) | 4 (22) | 19 (32) | 2 (40) | 2 (9) | 10 (22) |
| 5 or more | 8 (5) | 0 (0) | 5 (9) | 0 (0) | 1 (4) | 2 (4) |
| COVID-19 Antiviral Drug Use at Baseline<br>(Between 90 days prior to and 7 days after first visit) |  |  |  |  |  |  |
| No | 39 (26) | 3 (17) | 11 (19) | 2 (40) | 4 (17) | 19 (42) |
| Yes | 111 (74) | 15 (83) | 48 (81) | 3 (60) | 19 (83) | 26 (58) |
| 88 |  |  |  |  |  |  |
| Viral Lineage |  |  |  |  |  |  |
| BA.1 | 4 (3) | 2 (11) | 1 (2) | 1 (20) | 0 (0) | 0 (0) |
| BA.2 | 46 (31) | 9 (50) | 19 (32) | 1 (20) | 8 (35) | 9 (20) |
| BA.4 | 9 (6) | 2 (11) | 1 (2) | 0 (0) | 2 (9) | 4 (9) |
| BA.5 | 43 (29) | 2 (11) | 17 (29) | 2 (40) | 8 (35) | 14 (31) |
| Unknown | 48 (32) | 3 (17) | 21 (36) | 1 (20) | 5 (22) | 18 (40) |
| Days to last SARS-CoV-2-positive RT-PCR test result |  |  |  |  |  |  |
| Median days (IQR) | 9 (2-26) | 11 (3-44) | 16 (4-29) | 32 (20-33) | 9 (3-27) | 4 (0-9) |

**Abbreviations:** SARS-CoV-2 = severe acquired respiratory syndrome coronavirus 2; SOT/HSCT = solid organ transplant/hematopoietic stem cell transplant; AIDS = acquired immunodeficiency syndrome; IQR = interquartile range; rRT-PCR = real time reverse transcription polymerase chain reaction
\*Participants were enrolled from the following medical centers in 5 U.S. states: Michigan Medicine (Ann Arbor, MI), Vanderbilt University Medical Center (Nashville, TN), Montefiore Medical Center (Bronx, NY), Hennepin County Medical Center (Minneapolis, MN), and Washington University Medical Center (St. Louis, MO).
#First positive SARS-CoV-2 test was used for asymptomatic patients ^Includes any COVID-19 vaccine formulation

All 150 patients were enrolled during the period of SARS-CoV-2 Omicron variant predominance. A lineage was determined for 102/150 (68%) patients, including 4/150 (3%) with BA.1, 46/150 (31%) with BA.2, 9/150 (6%) with BA.4, 43/150 (29%) with BA.5, and lineage was unknown for 48/150 (32%).

### Nucleic acid and culture positivity over time

All 150 patients were enrolled based on a positive SARS-CoV-2 rRT-PCR test obtained in the clinical setting. Specimens from the enrollment visit tested positive by rRT-PCR for SARS-CoV-2 at the central laboratory for 121/150 (81%) patients. Of these, the enrollment visit was the last positive SARS-CoV-2 rRT-PCR test at the central laboratory for 80/121 (66%) patients and only 41/121 (34%) patients had at least one follow-up visit. This included 28 patients with a positive test at 1 follow-up visit, 7 patients with a positive test at 2 follow-up visits, and 6 patients with a positive test at ≥3 follow-up visits.

The individual trajectories of total viral RNA (as estimated from rRT-PCR Ct value) and the time to last positive test varied significantly across immunosuppressed groups (Figure 1). The median time to last positive rRT-PCR test overall was 9 days (IQR 2-26); the AIDS group had the longest median time to last positive rRT-PCR test (32 days, IQR 20-33), followed by the SOT/HSCT group (16 days, IQR 4-29) and B cell dysfunction group (11 days, IQR 3-44). The autoimmune/autoinflammatory group had the shortest time to last positive test at 4 days (IQR 0-9). Compared to the autoimmune/autoinflammatory group, patients in the B cell dysfunction group (aHR 0.32, 95% CI 0.15-0.64), SOT/HSCT group (aHR 0.60, 95% CI 0.38-0.94), and AIDS group (aHR 0.28, 95% CI 0.08-1.00) had longer duration of infection, defined as time to last positive rRT-PCR test. No other covariates, including age, sex, ethnicity, vaccination status, or baseline antiviral use were associated with duration of infection (Supplemental Table 3).

**Figure 1.**
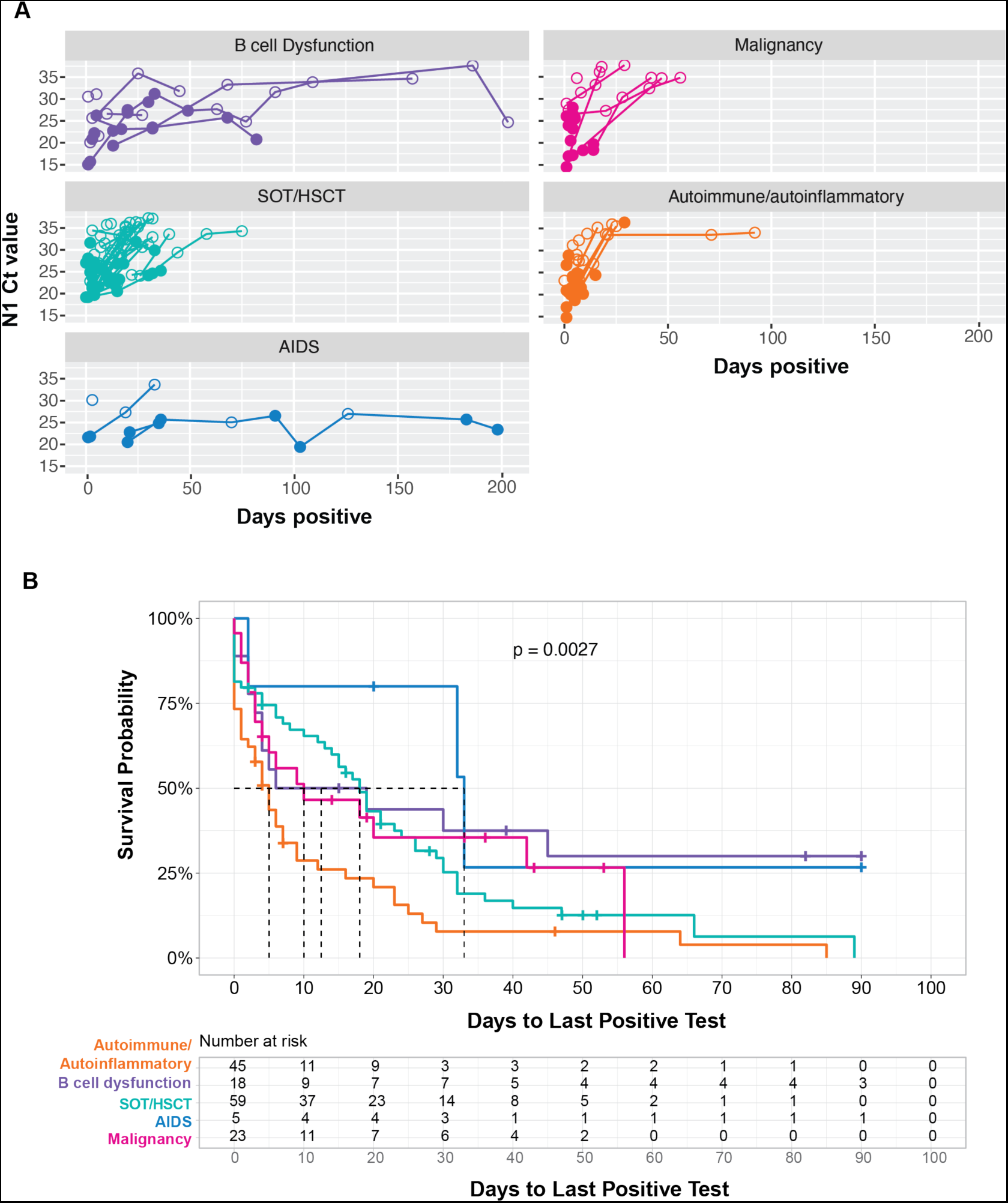
Temporal dynamics of SARS-CoV-2 RNA viral load and culture positivity in 121 immunocompromised patients with SARS-CoV-2 infection. (A) Cycle threshold (Ct) values for total SARS-CoV-2 RNA and virus culture isolation over time in 121 patients by immunocompromised group. Open and closed circles indicate culture negative and positive specimens, respectively. (B) Kaplan-Meier survival curves showing time to last positive rRT-PCR test by immunocompromised group. p = 0.003 for difference in time to last positive specimen across groups.

Ct values for subgenomic viral RNA, a marker of active viral replication [32,41], tracked with total viral RNA Ct values across patients and timepoints (Supplemental Figure 2). Of the 192 specimens positive for SARS-CoV-2 by rRT-PCR, 93 (48%) yielded positive viral culture. A positive culture for SARS-CoV-2 was achieved in 65% of specimens with a total viral RNA Ct ≤32 and 4% with Ct >32. Thirty-eight (25%) patients were positive for SARS-CoV-2 by rRT-PCR ≥21 days; of these, 16 (11%) patients had ≥2 sequenced specimens with Ct ≤32 over ≥21 days (Supplemental Figure 3). Of these, 5 exhibited very prolonged replication for >56 days, including one patient with SARS-CoV-2 positivity by rRT-PCR for 207 days and by culture for 198 days, a second patient with SARS-CoV-2 positivity by rRT-PCR for 82 days and by culture for 82 days, a third patient with SARS-CoV-2 positivity by rRT-PCR for 157 days and by culture for 32 days, a fourth patient with SARS-CoV-2 positivity by rRT-PCR for 75 days and by culture for 30 days, and a fifth patient with SARS-CoV-2 positivity by rRT-PCR for 203 days and by culture for 49 days (Supplemental Table 2).

### Evolutionary divergence in persistent infection

We obtained high depth of coverage sequence data suitable for identifying the whole genome consensus and iSNV from 149 (78%) specimens from 104 patients (Supplemental Figure 4). To account for the large number of fixed Omicron-defining mutations present in these samples, any mutation present at ≥98% frequency in the first sample for each patient was considered an Omicron-related mutation and was not examined further (see Methods). Using this definition, 93 patients had *de novo* mutations or iSNV, and 65 of these had *de novo* non-synonymous mutations. There was no relationship between the number of iSNV identified and total viral RNA Ct value (Supplemental Figure 5). At each time point we identified similar numbers of iSNV, consistent with the dynamic gain and loss of both nonsynonymous and synonymous mutations (Figure 2A). We found evidence for significant divergence in multiple genes, including ORF1a, ORF1b, and spike (Supplemental Table 4). At a genome level, patients with persistent infection lasting ≥21 days (n=16), compared to patients with short-term infection <21 days (n=72), had an increased nonsynonymous divergence rate (2.73×10^-6^ vs. 5.75×10^-7^ per site per day, Mann Whitney U test p=0.03) and similar synonymous divergence rate (Mann Whitney U test p=0.29). The overall mutation rate (including both non-synonymous and synonymous mutations) was similar between patients with short-term and persistent infection (5.80 x 10^-6^ and 3.95 x 10^-6^, respectively; Mann Whitney U test p=0.16) (Figure 2B).

**Figure 2.**
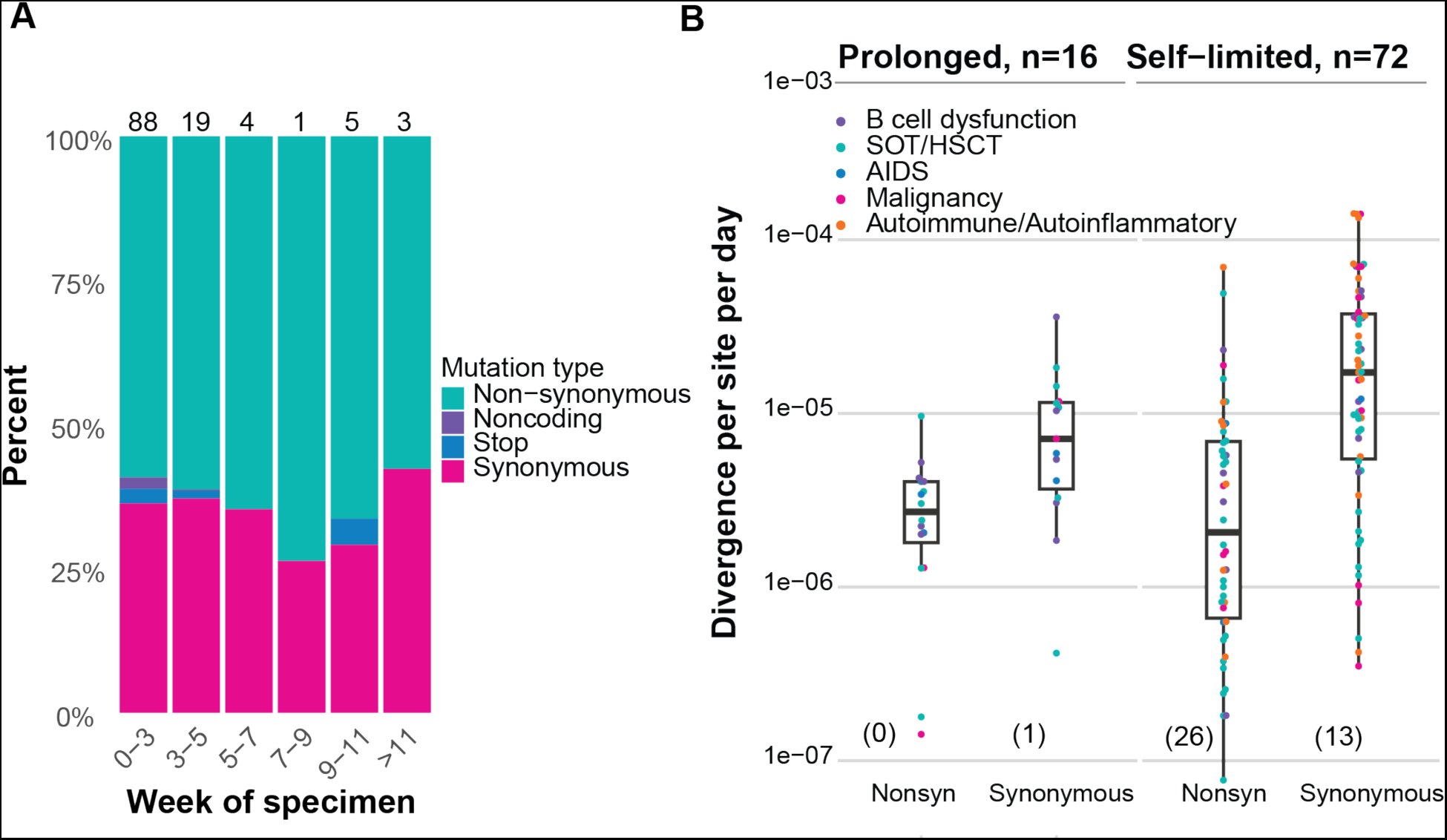
Within-host evolution of SARS-CoV-2 in 104 immunocompromised patients. (A) Stacked columns show percent of newly arising mutations identified at >2% frequency for specimens collected during the indicated time periods with mutation types color-coded: nonsynonymous (teal), noncoding (purple), stop codon (blue), synonymous (pink). Number of samples in each group is listed atop each bar; n = 93. (B) Genome-wide within-host divergence rate for individuals positive for SARS-CoV-2 by rRT-PCR for <21 days (n = 72) compared to those positive for ≥21 days (n = 16). Individuals in each group with rates of zero (e.g., no mutations identified ≥2% frequency in the final specimen) are not plotted given log transformation of y-axis and are indicated in parentheses at bottom of plot; however, these were included in the statistical analysis. Mann Whitney U test p=0.03 for differences in nonsynonymous rates and p=0.29 for differences in synonymous rates between prolonged and self-limited infections. Points are color-coded by immunocompromised group: B cell dysfunction, purple; SOT or HSCT, teal; AIDS, blue; non-B cell malignancy, pink; autoimmune/autoinflammatory, orange.

### Shared and within-host mutational evolution

We examined if any newly arising mutations were shared among multiple patients, which would provide evidence for positive selection [42–44]. There were very few shared mutations in the study population (Figure 3A). The K444N substitution, in the receptor binding domain of Spike, was shared by 9 patients. This mutation has been associated with monoclonal antibody resistance, and 8 patients with this mutation received monoclonals [45,46]. The T1542I and T4311I substitutions, both in ORF1a and each shared by 4 patients (eventually achieving dominance in one patient), have not previously been reported in the literature and peaked at <1% frequency in the global population [47,48]. Five patients had new insertions or deletions at consensus level, most of which were not shared among multiple patients. Four deletions were shared by two patients each, all in the spike N-terminal domain: L141, G142, V143, and Y144.

**Figure 3.**
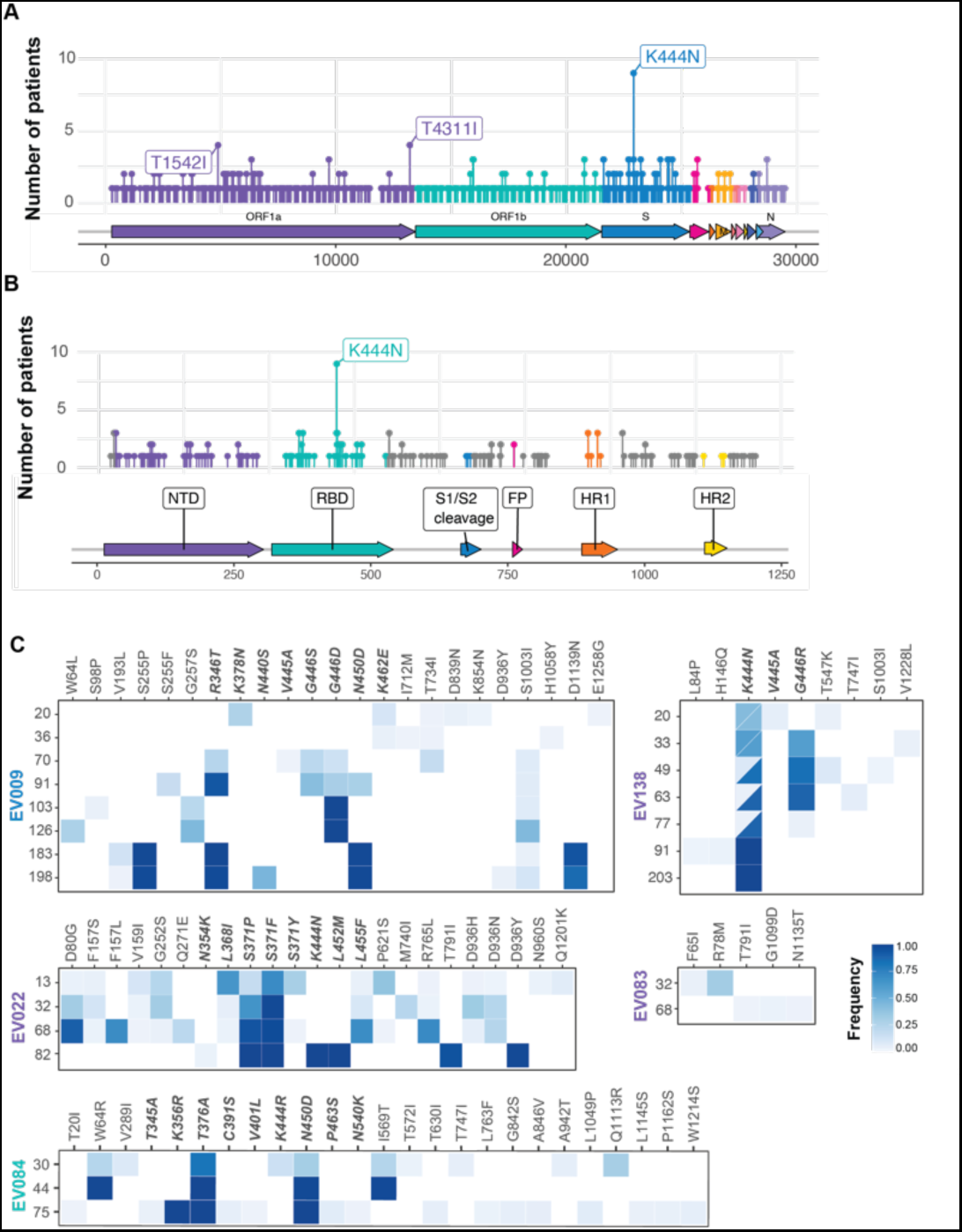
*De novo* non-synonymous SARS-CoV-2 mutations in 65 immunocompromised hosts. (A) Mutations shared by the indicated number of individuals (y-axis), color coded by gene. Amino acid substitutions are labeled if shared by ≥5% (n=4) of patients. (B) Mutations in spike shared by the indicated number of individuals (y-axis), color coded by domain. Amino acid substitutions are labeled if shared by ≥5% (n=4) of patients. (C) Heatmaps of *de novo* nonsynonymous mutations in SARS-CoV-2 spike and their frequencies in five individuals with infections lasting >56 days and with ≥2 sequenced samples. Patients are color coded by immunocompromised group: B cell dysfunction, purple; SOT or HSCT, teal; AIDS, blue; non-B cell malignancy, pink; autoimmune/autoinflammatory, orange. Day of infection is indicated on the Y axis. EV138 received bebtelovimab on day 1, and EV022 received bebtelovimab at day 68. Mutations in the receptor binding domain are indicated by bold italics. Bisected squares indicate more than one codon mutation identified produced the same amino acid substitution. In EV022, shading at position 371 reflects the combined frequency of the 371F and 371P alleles.

Of the 5 patients with very prolonged viral shedding, 4 accumulated consensus level mutations in spike, 61% of which were in the receptor binding domain (RBD, Figure 3C, Supplemental Table 5). As in the entire patient population, there were few shared mutations. The K444N, G446D/R, and N450D substitutions were the only mutations shared by multiple patients; all are associated with monoclonal antibody resistance, but only two of the five patients received a monoclonal antibody. None of these mutations have been prevalent globally; K444N peaked at 2% global frequency, and N450D at 3% global frequency, both in November 2022.

Of 23 consensus spike mutations identified in these five individuals, most have been seen in subsequent Omicron lineages. The 5 mutations (F157L, R346T, L368I, S371F, and T376A) that subsequently achieved >10% frequency globally were seen only in individual patients and not shared. The R346T substitution in one patient, which was not characteristic of the infecting BA.2.12.1 lineage, was subsequently a defining mutation in XBB and BQ.1.1 lineages. The L368I and 371F substitutions, which were not characteristic of the infecting BA.1.1 lineage, were seen in later Omicron lineages (371F) and XBB (L368I) lineages. Both were present at >60% frequency in the patient’s first sample (day 13 of infection), making them less remarkable as markers of within-host evolution fostered by persistent infection. Notably, mutations at K356, V445, G446, and N450 –– all identified in these patients but not frequently in the general population – are mutated in the recently identified and highly divergent BA.2.86 variant under monitoring.

Neutralization assays with pooled sera against the initial and evolved viruses from patients with prolonged infection indicated that the evolved virus from patient EV138 (de novo spike mutations K444N, G446R) was antigenically distinct (mean ± sd FRNT50 fold change -3.26 ± 1.02, vs. matched initial virus, n=6 serum pools, Supplemental Table 6) while the evolved virus from patient EV022 (de novo spike mutations K444N, L452M) was not (FRNT50 -0.25 ±. 1.09 fold change vs. matched initial virus).

### Impact of antiviral treatment

We examined mutational patterns in patients with pre-treatment and at least one post-treatment sequenced sample to determine if any resistance mutations developed in our study population. A total of 115 (77%) patients received one or more antiviral treatments (including remdesivir, molnupiravir, nirmatrelvir/ritonavir, convalescent plasma, or any monoclonal antibody) at any time during the study period. Of the 42 patients who received a monoclonal antibody (including bebtelovimab, sotrovimab, and/or tixagevimab/cilgavimab), 16 (38%) had a post-treatment sample; of these, 10 (62%) had *de novo* (i.e., new in patient) nonsynonymous mutations in spike (Figure 4A, Table 2). There were several shared mutations among these patients, most of which were between positions 444-446 and have been associated with monoclonal antibody resistance [45,46].

**Figure 4.**
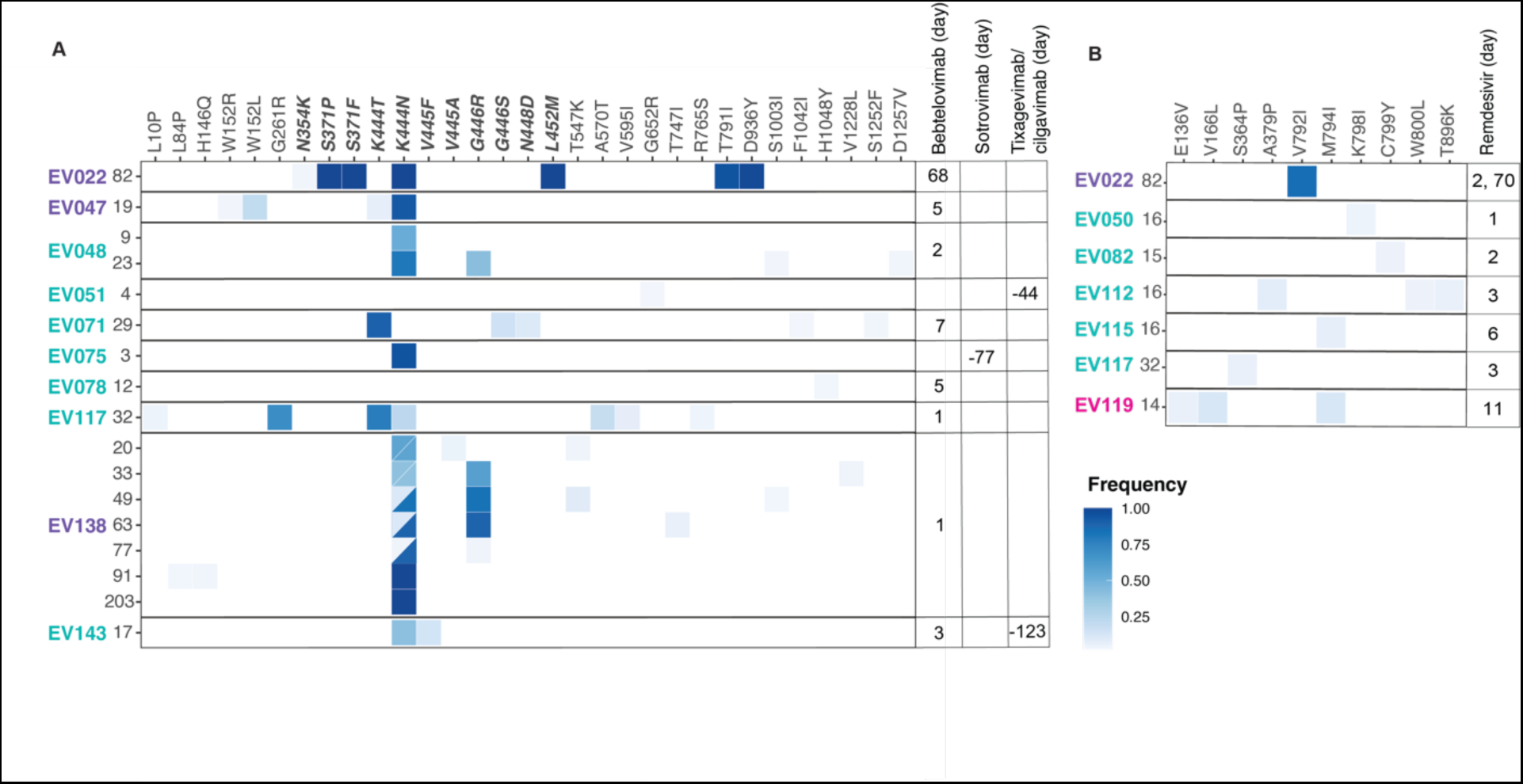
Mutations in 15 patients who received antiviral treatment. (A) Heatmap of *de novo* nonsynonymous mutations in SARS-CoV-2 spike among immunocompromised patients who received monoclonal antibody (bebtelovimab, sotrovimab, and/or tixagevimab/cilgavimab) and had a post-treatment sample that was sequenced (n=10). Sixteen patients had a post-treatment monoclonal antibody sample; of these, 10 had *de novo* non-synonymous mutations in spike. Patients are color coded by immunocompromised group: B cell dysfunction, purple; SOT or HSCT, teal; AIDS, blue; non-B cell malignancy, pink; autoimmune/autoinflammatory, orange. Monoclonal antibody received and treatment timepoints are denoted for each patient. Mutations in the receptor binding domain are shown in bold italics. Bisected squares indicate more than one codon mutation produced the same amino acid substitution. (B) Heatmap of mutations in SARS-CoV-2 nsp12 (RNA dependent RNA polymerase) among immunocompromised patients who received remdesivir and had a post-treatment sample (n=7). 17 patients had a post-treatment remdesivir sample; of these, 7 had *de novo* non-synonymous mutations in nsp12. In both (A) and (B), the day of infection is indicated to the left of heatmap, and the day of treatment is indicated to the right.

**Table 2.**
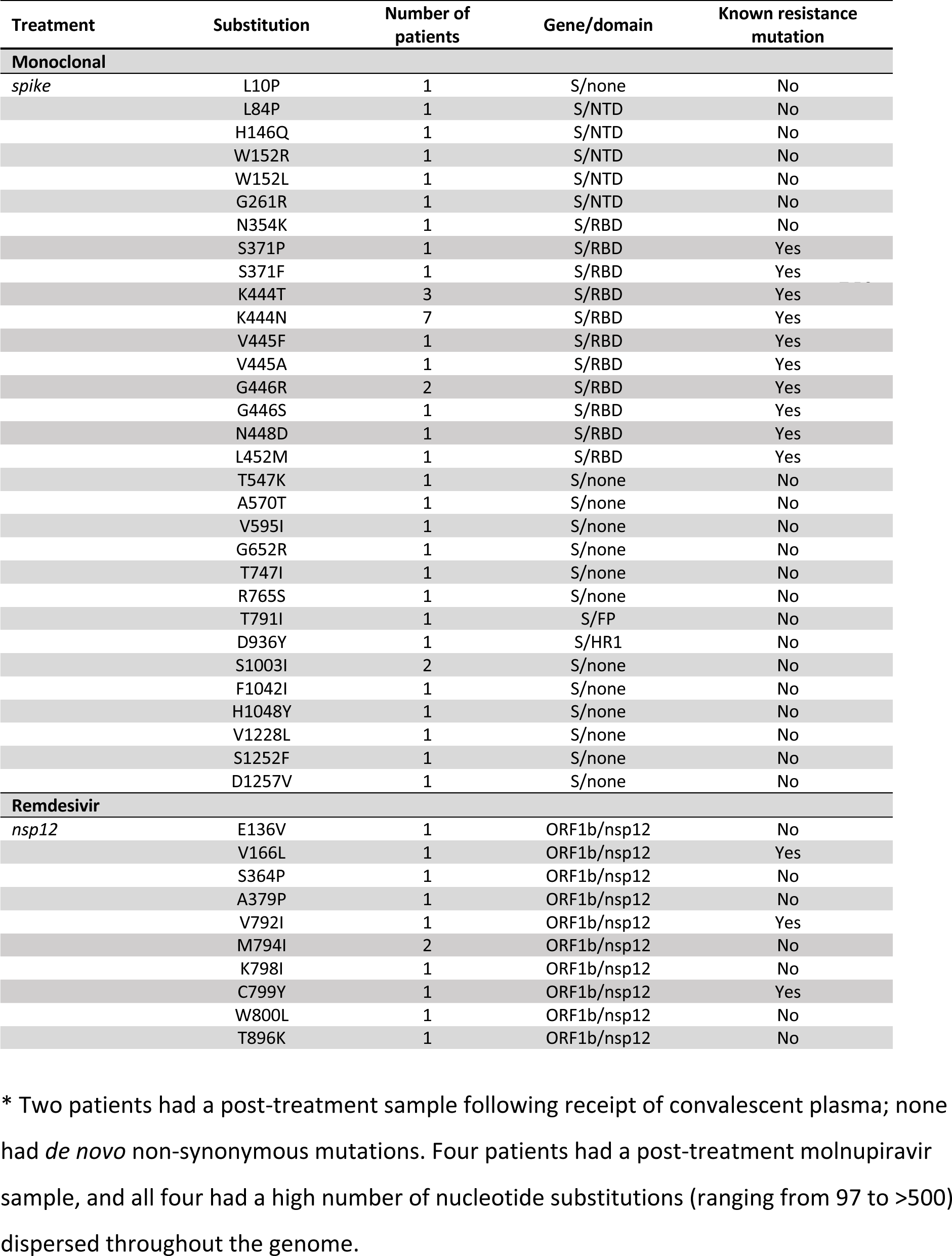
*De novo* mutations identified in immunocompromised patients after treatment with SARS-CoV-2 antivirals — IVY Network, 5 U.S. States, April 11, 2022 – February 1, 2023.

Among patients treated with antiviral drugs, 17 of 68 (25%) remdesivir-treated patients had at least one post-treatment sample; 7/17 (41%) had *de novo* mutations in nsp12, the target of remdesivir. Most of these mutations were present at very low frequency and were not shared among multiple patients; only one, M794I, was shared by 2 patients (Table 2, Figure 4B). While most have not been specifically associated with remdesivir resistance, three of these newly arising mutations have been associated with resistance, and several others are in close proximity to C799, where remdesivir resistance-associated substitutions have been identified [45,49]. Only one of the resistance-associated mutations, V792I, was present at high frequency in a single patient [6]. Five out of 20 (25%) nirmatrelvir/ritonavir-treated patients had a post-treatment sample, none of which had any mutations in nsp5 (Mpro) [45,50]. Five patients received molnupiravir, and 4 had a post-treatment sample. All four had a high number of nucleotide substitutions (ranging from 97 to >500), most of which were present at low frequency and distributed evenly across the genome; most of these specimens were viral culture negative.

Most patients who received antiviral treatments cleared their infections. Four who received any treatment (1 who received nirmatrelvir/ritonavir, 1 who received molnupiravir, 1 who received bebtelovimab, and 1 who received both bebtelovimab and remdesivir), went on to have very prolonged viral shedding >56 days (Supplemental Figure 6).

## DISCUSSION

In this prospective, multicenter analysis conducted during the Omicron period, prolonged replication-competent SARS-CoV-2 infection among a diverse group of patients with moderate to severe immunocompromise was uncommon. Additionally, while numerous case reports of SARS-CoV-2 infection in immunocompromised hosts have documented significant mutation accumulation in spike that mirrors mutational profiles in VOC, our analysis demonstrated comparatively restricted SARS-CoV-2 evolution over a wide spectrum of immunocompromising conditions. We found that the within-host rate of evolution in immunocompromised hosts – captured as divergence – was similar in short-term and long-term infection. Our data suggest that the main difference in some immunocompromised hosts is the length of the infectious period, which allows for mutation accumulation without the constraint of transmission. In the few individuals with prolonged infection, we found accumulation of mutations within the RBD; the most prominent alterations have rarely, if ever, been detected in subsequent SARS-CoV-2 sequences in global databases. We did find several substitutions also present in concurrent or subsequent Omicron lineages.

Our analysis identified B cell dysfunction/depletion due to lymphoma or myeloma, or anti-CD20/anti-CD19 therapy, as a strong risk factor for longer duration of SARS-CoV-2 infection among the immunocompromised population (see also [51]). This is consistent with published case reports, where nearly all of those with infections lasting >150 days had B cell malignancy with receipt of anti-CD20 antibodies or CAR-T cells [7,9,15–18,52–65]. Although we enrolled 59 SOT and HSCT patients with ongoing T cell immunosuppression – due to tacrolimus, prednisone, and/or mycophenolate – only one had infection lasting >56 days. These findings highlight the importance of antibodies in SARS-CoV-2 clearance, both consistent with what has been seen in other viruses, and with emerging evidence of antibodies as correlates of protection in SARS-CoV-2 [66–70]. One of our cases with very prolonged infection (>200 days) was in a person living with AIDS with a CD4 T cell count <50 and uncontrolled HIV replication. This is consistent with both case reports [17] and a systematic review [71], which found that prolonged replication in people living with HIV is seen at extremely low CD4 counts (<50) and high HIV viral loads. This may also reflect impaired humoral immunity, as B cell responses are compromised in advanced AIDS [72].

Because we collected specimens at regular intervals for rRT-PCR and viral culture, we were able to examine the kinetics of viral clearance in patients who received antiviral therapy. Among 115 individuals treated with antivirals, only four did not clear their infection. In the four failures, both patients who received bebtelovimab developed associated resistance mutations. A third patient received remdesivir and developed a mutation associated with remdesivir resistance, V792I. While our data suggest that most immunocompromised patients have good virological responses to antiviral treatment, there is a need for further studies of extended treatment courses or combination therapy in those at highest risk for prolonged infection [73,74].

The observed viral evolutionary dynamics in our surveillance cohort may differ from what was reported earlier in the pandemic. Many published cases of prolonged SARS-CoV-2 infection were in patients who had received convalescent plasma or early generation monoclonal antibodies (e.g., bamlanivimab), both of which tend to select for the same escape mutations (e.g., N-terminal domain mutations, E484K, and others) as infection-or vaccine-induced antibodies. In immunocompromised hosts treated with therapeutic antibodies, SARS-CoV-2 is likely to encounter intensified selective pressures similar to those at the global scale. In the absence of treatment, however, many immunocompromised hosts will have little antibody pressure on the spike protein, and the within-host and current global selective pressures may not align. People living with AIDS may differ from those with B cell dysfunction or depletion, as the former might plausibly mount an attenuated antibody response sufficient to select for mutations in the absence of viral eradication. While the origin of the highly divergent BA.2.86 variant is unclear, mutations in this variant at positions 445 and 446 were identified in 2/3 patients and 3/5 patients, respectively, who received monoclonal antibodies. Our serological data suggest that K444N and G446R together lead to increased neutralizing antibody escape.

Our study is subject to limitations. First, although we included 150 immunocompromised patients with SARS-CoV-2 infection in this longitudinal evaluation, only 41 (27%) had a follow-up specimen that tested positive by rRT-PCR, limiting the number of individuals in whom relevant features of viral evolution emerging on the population level could be assessed. Second, the definition of immunocompromise in the study was intentionally broad to capture as many patients as possible who might be at risk for prolonged infection and avoid bias toward any particular group. However, this breadth likely also led to the inclusion of patients with modest immune impairment who were less likely to experience prolonged infection and virus evolution. Third, the frequency of specimen collection at 2–4 week intervals was optimal to assess interval change in mutations among patients with prolonged infection, but too infrequent to determine precise estimates of the duration of rRT-PCR-positivity, as 73% did not have a positive specimen after enrollment. Fourth, we did not enroll immunocompetent patients who could have provided a referent group to compare duration of rRT-PCR-positivity by type of immunosuppression. Prior studies showing that prolonged positivity in immunocompetent adults is rare contributed to our decision not to include an immunocompetent comparator group [41]. Finally, our results from a US-based population may not generalize to immunocompromised hosts in other settings. Recipients of SOT, HSCT, CAR-T, and/or anti-CD20 monoclonal antibodies were over-represented in our study and people living with AIDS were under-represented. Findings may differ in locations where AIDS is more prevalent and access to SARS-CoV-2 antivirals and COVID-19 vaccines is lower [17].

In conclusion, in this prospective cohort of immunocompromised adults with SARS-CoV-2 infection, duration of infection and evolution of SARS-CoV-2 were observed more frequently in patients with B cell malignancy and B cell depletion. With extended viral replication, these immunocompromised individuals can accumulate significant numbers of mutations in the spike protein and elsewhere across the genome and exhibit marginally accelerated viral evolution. In our cohort of individuals infected with Omicron variant SARS-CoV-2, mutations arising within immunocompromised hosts were only weakly predictive of subsequent Omicron mutations at a population scale, suggesting that alternative genomic surveillance approaches may be more useful [75]

## Data Availability

All data produced in the present work are contained in the manuscript. Raw sequencing reads are available on the NCBI short read archive under BioProject PRJNA896930 and consensus sequences are available on GISAID.

## ACKNOWLEDGEMENTS

We thank all participants in this study and all contributors to the Global Initiative on Sharing All Influenza Data (GISAID) sequence database.

## FUNDING ACKNOWLEDGEMENT

Primary funding for this study was provided by the US Centers for Disease Control and Prevention (CDC) (award 75D30121F00002). Scientists from the US CDC participated in all aspects of this study, including its design, analysis, interpretation of data, writing the report, and the decision to submit the article for publication.

ZR was supported by NIH T32HL007749.

## DISCLAIMER

The findings and conclusions in this report are those of the authors and do not necessarily represent the official position of the Centers for Disease Control and Prevention (CDC).

---

**Supplemental Table 1.** List of qualifying medications that are immunosuppressive, immunomodulatory, or myelosuppressive (see attached)

**Supplemental Table 2.** Clinical, demographic, viral, and specimen data for enrolled patients (see attached)

**Supplemental Table 3.** Output of Cox proportional hazards model for time to last positive SARS-CoV-2 rRT-PCR test.

|  | Hazard Ratio | Lower 95% CI | Upper 95% CI |
| --- | --- | --- | --- |
| B cell dysfunction vs. Autoimmune/Autoinflammatory | 0.315 | 0.154 | 0.644 |
| Post-SOT/HSCT vs. Autoimmune/Autoinflammatory | 0.597 | 0.38 | 0.939 |
| AIDS vs. Autoimmune/Autoinflammatory | 0.282 | 0.08 | 0.996 |
| Malignancy vs. Autoimmune/Autoinflammatory | 0.582 | 0.312 | 1.085 |
| Age | 0.994 | 0.981 | 1.008 |
| Sex - Male vs. Female | 1.035 | 0.7 | 1.529 |
| Black vs. White | 0.852 | 0.55 | 1.32 |
| Hispanic vs. White | 2.049 | 1.002 | 4.191 |
| Other / Unknown vs. White | 0.671 | 0.269 | 1.672 |
| One or More Dose of Vaccine vs. Not Vaccinated | 1.653 | 0.775 | 3.527 |
| Baseline Antiviral Use - Yes vs. No | 0.878 | 0.572 | 1.35 |

\* Variables included in the final model were immunocompromised group, age, sex, race, vaccination status, and receipt of antiviral drug at baseline (see Methods).

**Supplemental Table 4.** Within-host divergence rates by gene for synonymous, nonsynonymous, and stop-codon mutations.

| Gene | Mutation type | Zero count (%) | Median divergence rate | IQR | p-value* |
| --- | --- | --- | --- | --- | --- |
| **ORF1a** | Non-synonymous | 51 (58%) | 0 | 1.34E-06 | 3.56E-06 |
|  | Stop | 85 (97%) | 0 | 0 | 1 |
|  | Synonymous | 37 (42%) | 1.89E-06 | 2.24E-05 | 7.37E-09 |
| **ORF1b** | Non-synonymous | 47 (53%) | 0 | 1.82E-06 | 7.55E-07 |
|  | Stop | 87 (99%) | 0 | 0 | 1 |
|  | Synonymous | 59 (67%) | 0 | 1.22E-06 | 8.11E-05 |
| **S** | Non-synonymous | 51 (58%) | 0 | 8.94E-06 | 3.56E-06 |
|  | Stop | 87 (99%) | 0 | 0 | 1 |
|  | Synonymous | 53 (60%) | 0 | 4.07E-05 | 7.76E-06 |
| **ORF3a** | Non-synonymous | 82 (93%) | 0 | 0 | 1 |
|  | Stop | 86 (98%) | 0 | 0 | 1 |
|  | Synonymous | 82 (93%) | 0 | 0 | 1 |
| **E** | Non-synonymous | 87 (99%) | 0 | 0 | 1 |
|  | Stop | 88 (100%) | 0 | 0 | NA |
|  | Synonymous | 86 (98%) | 0 | 0 | 1 |
| **M** | Non-synonymous | 77 (88%) | 0 | 0 | 0.11571878 |
|  | Stop | 88 (100%) | 0 | 0 | NA |
|  | Synonymous | 83 (94%) | 0 | 0 | 1 |
| **ORF6** | Non-synonymous | 85 (97%) | 0 | 0 | 1 |
|  | Stop | 88 (100%) | 0 | 0 | NA |
|  | Synonymous | 85 (97%) | 0 | 0 | 1 |
| **ORF7a** | Non-synonymous | 82 (93%) | 0 | 0 | 1 |
|  | Stop | 88 (100%) | 0 | 0 | NA |
|  | Synonymous | 87 (99%) | 0 | 0 | 1 |
| **ORF8** | Non-synonymous | 81 (92%) | 0 | 0 | 0.67482814 |
|  | Stop | 85 (97%) | 0 | 0 | 1 |
|  | Synonymous | 88 (100%) | 0 | 0 | NA |
| **N** | Non-synonymous | 75 (85%) | 0 | 0 | 0.04985083 |
|  | Stop | 87 (99%) | 0 | 0 | 1 |
|  | Synonymous | 82 (93%) | 0 | 0 | 1 |

\* Wilcoxon signed rank test for difference in divergence rate (per site per day) vs. zero, Bonferroni adjusted p-value

**Supplemental Table 5.**
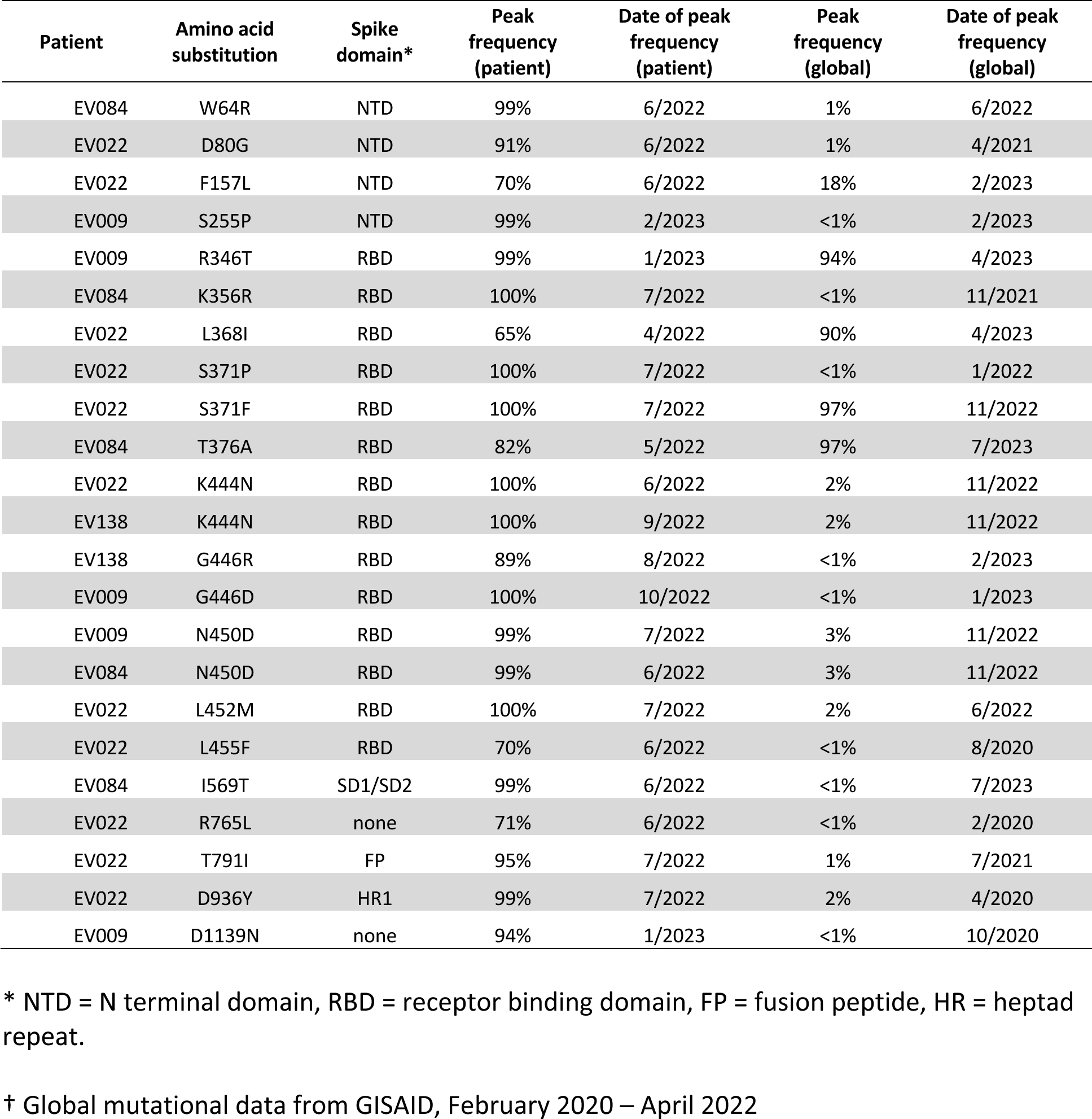
Consensus mutations in SARS-CoV-2 spike among 5 immunocompromised patients with ≥2 sequenced specimens and >56 days of RT-PCR positivity.

**Supplemental Table 6.** Neutralization FRNT50 of patient-derived viruses with serum pools.

| Patient and Sample Day | New Spike Mutations * | Pre-Vax Pool 1<sup>†</sup> | Post-Vax Pool 1<sup>†</sup> | Pre-Vax Pool 2<sup>†</sup> | Post-Vax Pool 2<sup>†</sup> | Pre-Vax Pool 3<sup>†</sup> | Post-Vax Pool 3<sup>†</sup> |
| --- | --- | --- | --- | --- | --- | --- | --- |
| EV138 day 20 (19 days post-bebtelovimab) | — | 68 | 166 | 38 | 278 | 574 | 992 |
| EV138 day 63 (62 days post-bebtelovimab) | K444N, G446R | 16 | 45 | 23 | 114 | 137 | 302 |
| *Fold Change* # | — | -4.25 | -3.7 | -1.7 | -2.4 | -4.2 | -3.3 |
| EV009 day 91 | R346T | 103 | 111 | 190 | 13 | 47 | 44 |
| *Fold Change* # |  | ND | ND | ND | ND | ND | ND |
| EV022 day 13 | — | 15 | 45 | 44 | 92 | 91 | 230 |
| EV022 day 82 (14 days post-bebtelovimab) | K444N, L452M | 13 | 33 | 53 | 114 | 63 | 253 |
| *Fold Change* # | — | -1.2 | -1.4 | 0.8 | 0.8 | -1.4 | 0.9 |

Abbreviations: FRNT, Focus reduction neutralization test
\* New mutations at >50% frequency relative to the initial sample from the same patient
<sup>†</sup> Three pools of matched pre- and post-bivalent vaccination sera from xx individuals (pool 1), xx individuals (pool 2), and xx individuals (pool 3)

**Supplemental Figure 1.**
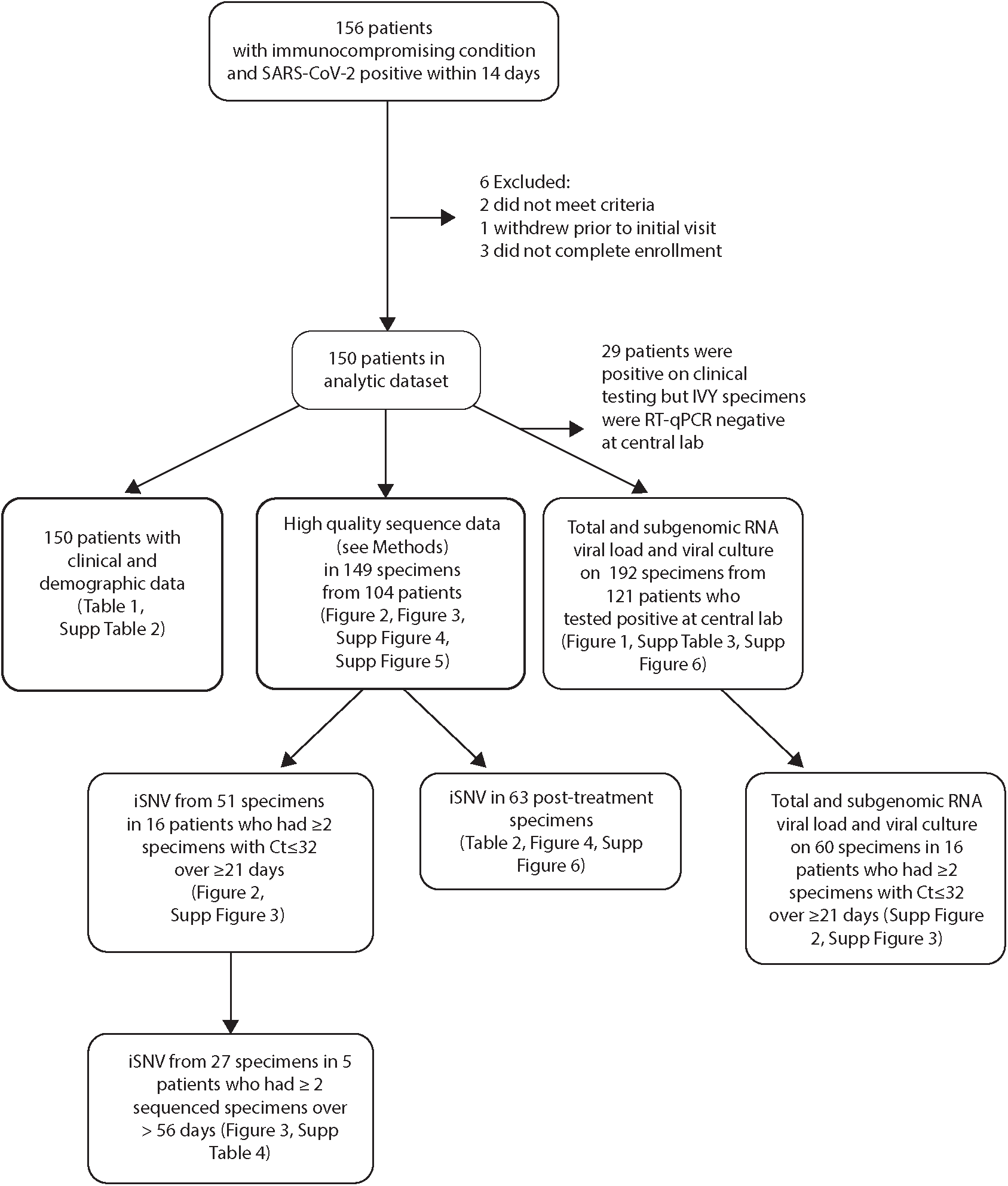
Flow diagram of enrolled patients indicating patients and data included in each analysis. iSNV = intrahost single nucleotide variants.

**Supplemental Figure 2.**
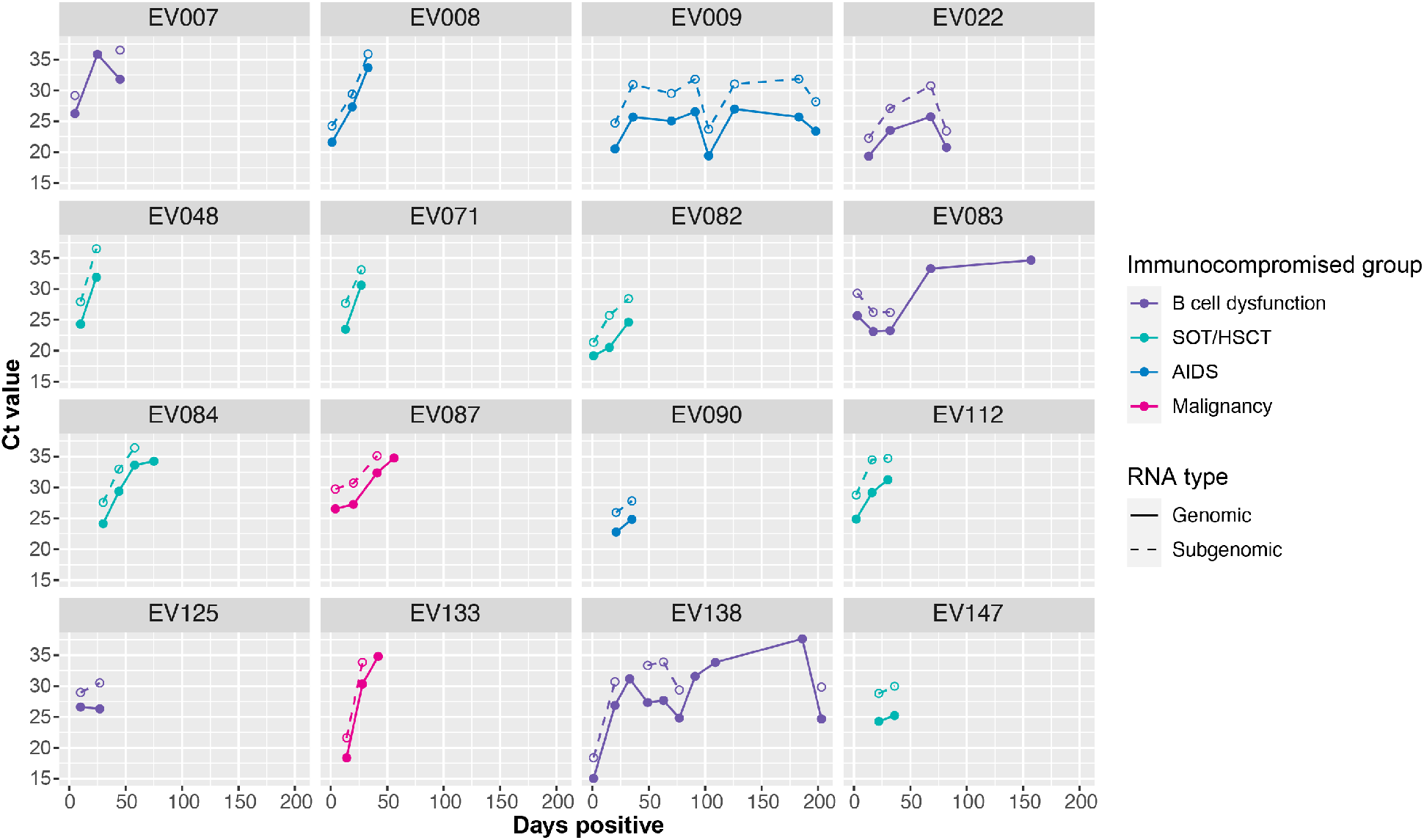
Plots show total (solid line, closed circles) and subgenomic N (dotted line, open circles) RNA viral load in serial specimens (day of infection, x-axis) for each of 16 immunocompromised patients who had detectable viral RNA in ≥2 specimens spanning ≥ 21 days. Lines and points are color coded by immunocompromised group: B cell dysfunction, purple; solid organ transplant (SOT) or hematopoietic stem cell transplant (HSCT), teal; AIDS, blue; non-B cell malignancy, pink; autoimmune/autoinflammatory, orange.

**Supplemental Figure 3.**
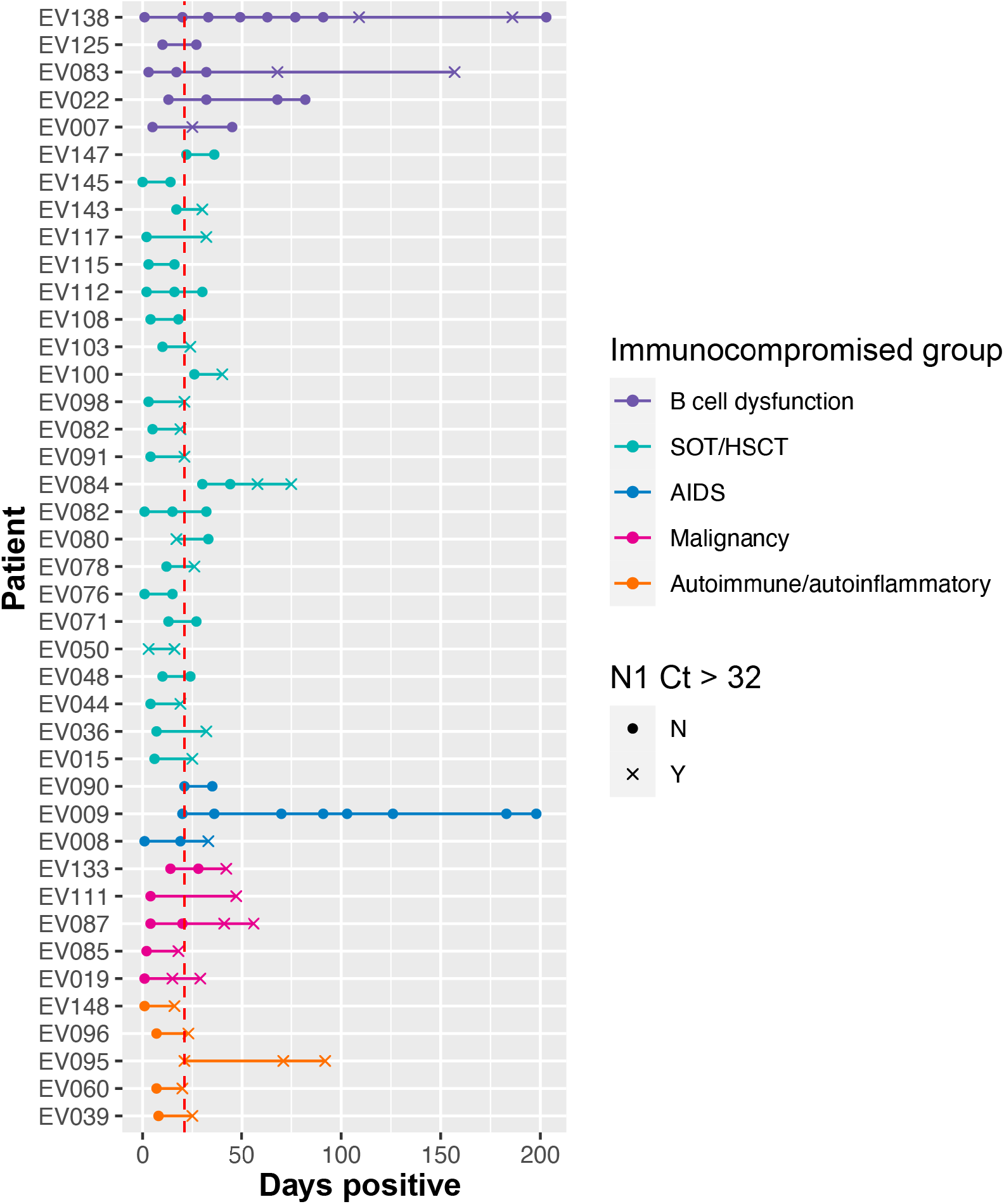
Diagram identifying 16 patients with prolonged infection; defined as patients who had ≥2 sequenced specimens with rRT-PCR Ct ≤32 spanning ≥21 days. Lines and points are color-coded by immunocompromised group: B cell dysfunction, purple; solid organ transplant (SOT) or hematopoietic stem cell transplant (HSCT), teal; AIDS, blue; non-B cell malignancy, pink; autoimmune/autoinflammatory, orange. Filled circles indicate specimens with rRT-PCR Ct ≤32 and x indicate specimens with rRT-PCR Ct >32.

**Supplemental Figure 4.**
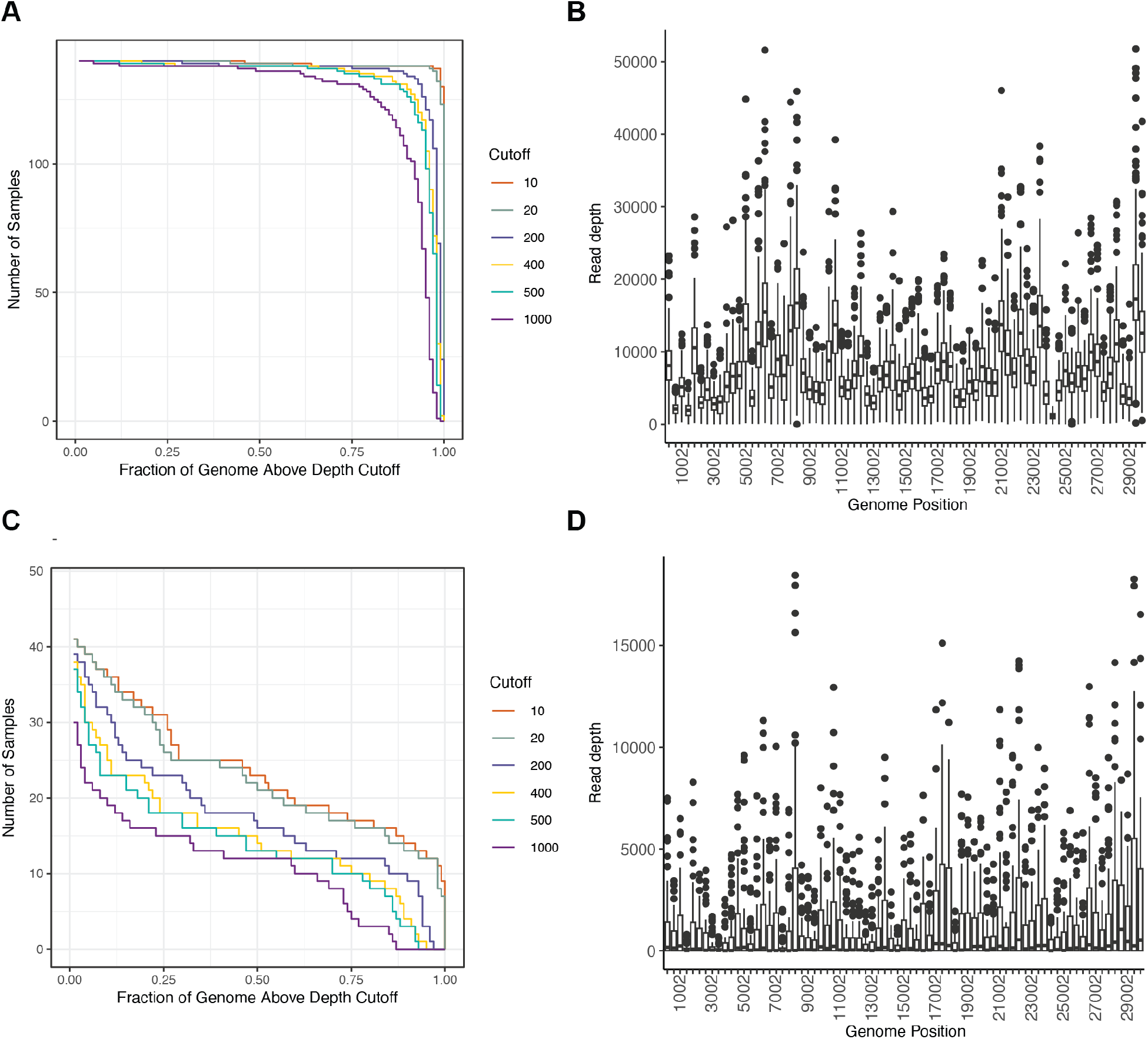
Depth of coverage in sequenced specimens. Plots showing (A) number of specimens (y-axis) with fraction of the genome (x-axis) covered at the indicated depths (lines, legend) and (B) read depth by genomic position (sliding window) for 140 specimens with rRT-PCR Ct ≤32. Plots showing (C) number of specimens (y-axis) with fraction of the genome (x-axis) covered at the indicated depths (lines, legend) and (D) read depth by genomic position (sliding window) for 52 specimens with rRT-PCR Ct >32.

**Supplemental Figure 5.**
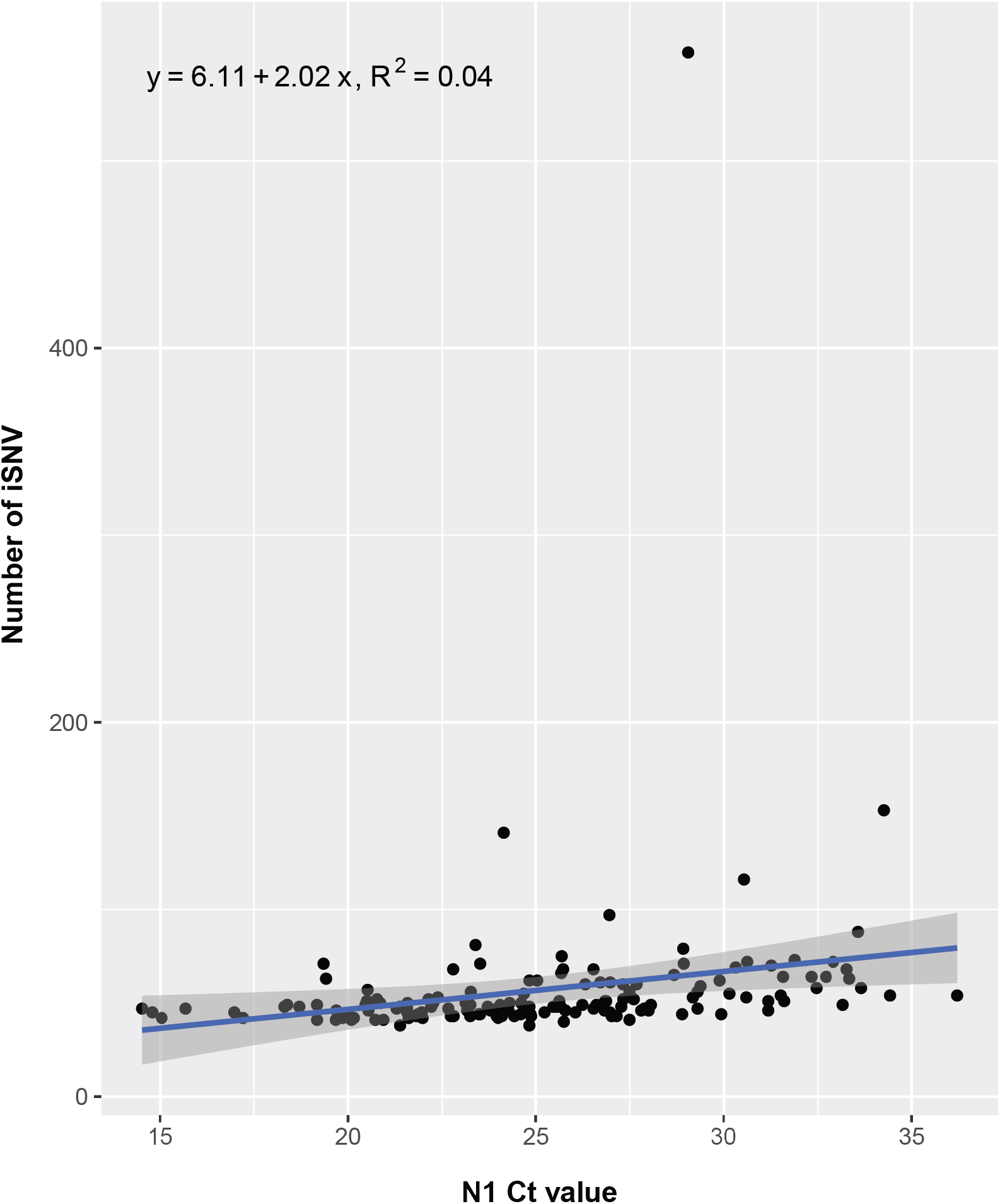
Number of intrahost single nucleotide variants (iSNV) identified at 2- 100% frequency (y-axis) per specimen (point) as a function of total SARS-CoV-2 RNA rRT-PCR Ct value. The regression line (blue), 95% confidence interval for the regression (shaded area), and the equation and R^2^ for the regression are indicated. The outlier point with over 500 iSNV represents a single specimen from a patient previously treated with molnupiravir.

**Supplemental Figure 6.**
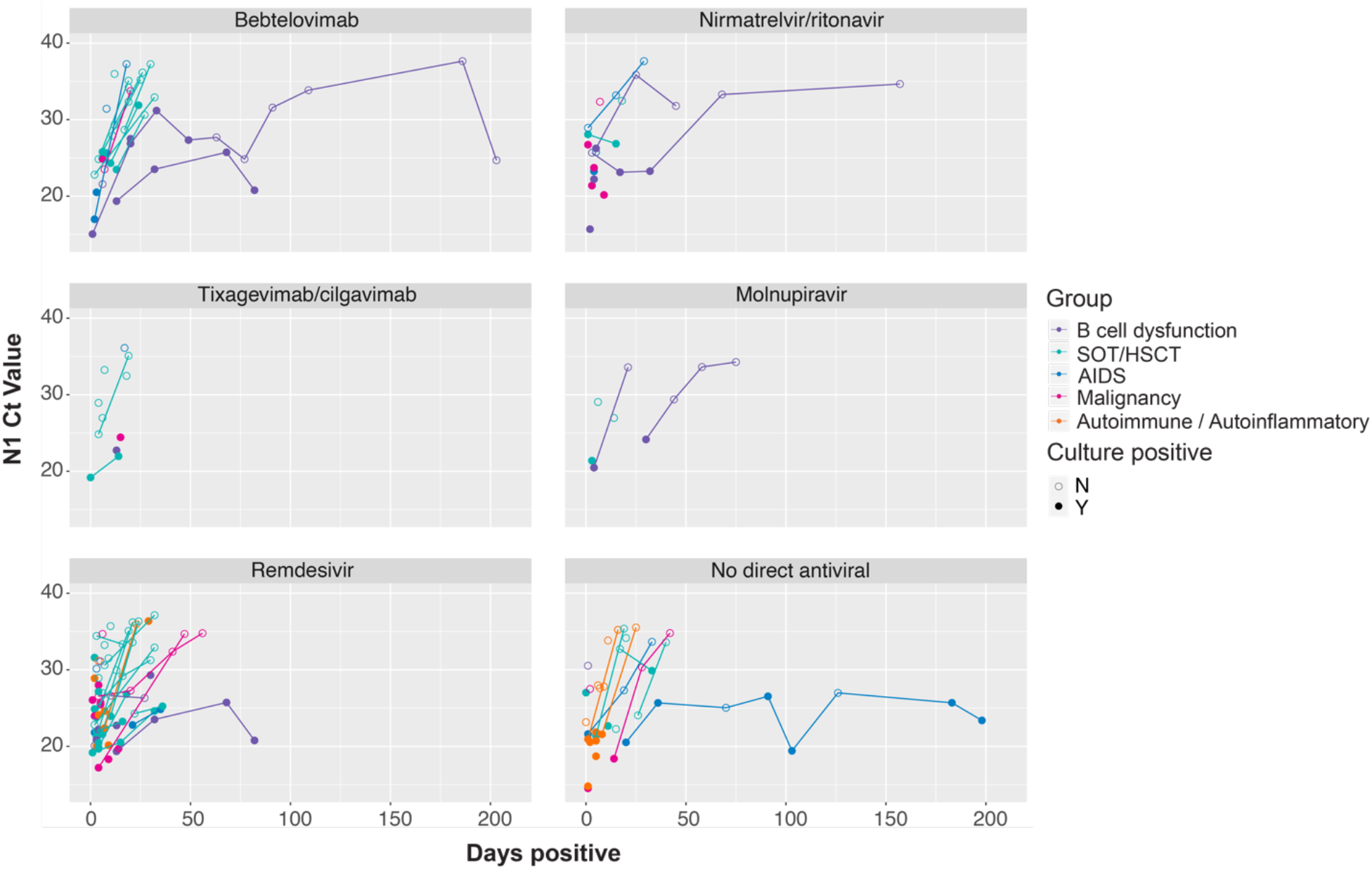
Plot of SARS-CoV-2 total RNA rRT-PCR Ct values for the 115 patients receiving indicated antiviral treatments. Lines and points are color-coded by immunocompromised group: B cell dysfunction, purple; solid organ transplant (SOT) or hematopoietic stem cell transplant (HSCT), teal; AIDS, blue; non-B cell malignancy, pink; autoimmune/autoinflammatory, orange.

