## Supplemental Table 1 for "SARS-CoV-2 shedding and evolution in immunocompromised hosts during the Omicron period: a multicenter prospective analysis"

| Generic Name | Trade Name |
| --- | --- |
| Abemaciclib | Verzenio |
| Adalimumab | Humira |
| Acalabrutinib | Calquence |
| Alefacept | Amevive |
| Amifostine | Ethyol |
| Anakinra | Kineret |
| Arsenic Trioxide | Trisenox |
| Asparaginase | Elspar |
| Auranofin | Ridaura |
| Aurothioglucose | Solganal |
| Axicabtagene ciloleucel | Yescarta |
| Azacitidine | Vidaza |
| Azathioprine | Imuran, Azasan |
| Basiliximab | Simulect |
| Belatacept | Nulojix |
| Belimumab | Benlysta |
| Bendamustine hydrochloride | Bendeka |
| Bevacizumab | Avastin |
| Bexarotene | Targretin |
| Bortezomib | Velcade |
| Brentuximab Vedotin | Adcetris |
| Brexucabtagene autoleucel | Tecartus |
| Budesonide | Multiple |
| Busulfan | Busulfex |
| Cabazitaxel | Jevtana |
| Capecitabine | Xeloda |
| Carboplatin | Paraplatin |
| Carfilzomib | Kyprolis |
| Carmustine | BiCNU |
| Certolizumab Pegol | Cimzia |
| Ciltacabtagene autoleucel | Carvykti |
| Cisplatin | Platinol, Platinol-AQ, CDDP |
| Cladribine | Mavenclad |
| Clofarabine | Clolar |
| Cyclophosphamide | Procytox |
| Cyclosporine | Gengraf; Neoral; SandIMMUNE |
| Dabrafenib | Tafinlar |
| Generic Name | Trade Name |
| Dacarbazine | DTIC-Dome; DTIC; DIC; imidazole carboxamide |
| Daclizumab | Zenapax |
| Dactinomycin | Cosmegen |
| Daratumumab | Darzalex |
| Dasatinib | Sprycel |
| Daunorubicin Citrate Liposome | Vyxeos |
| Decitabine | Inqovi; Dacogen; Demylocan |
| Denileukin Diftitox | Ontak |
| Denosumab | Prolia; Xgeva |
| Dexamethasone | Multiple |
| Docetaxel | Taxtere |
| Doxorubicin | Adriamycin |
| Eculizumab | Soliris |
| Efalizumab | Raptiva |
| Etanercept | Enbrel; Brenzys; Erelzi |
| Etoposide | Toposar |
| Everolimus | Afinitor; Afinitor Disperz; Zortress |
| Floxuridine | FUDR |
| Gefitinib | Iressa |
| Gemcitabine | Gemzar |
| Gold Sodium Thiomalate | Myochrysine |
| Golimumab | Simponi; Simponi Aria |
| Hydroxychloroquine | Plaquenil |
| Ibrutinib | Imbruvica |
| Idecabtagene vicleucel | Abecma |
| Ifosfamide | Ifex |
| Imatinib | Gleevec, Glivec |
| Infliximab | Avsola; Inflectra; Remicade; Remsima SC; Renflexis |
| Interferon Alfacon-1 | Alferon |
| Ipilimumab | Yervoy |
| Ixabepilone | Ixempra Kit |
| Ixazomib | Ninlaro |
| Leflunomide | Arava |
| Lenalidomide | Revlimid |
| Lisocabtagene maraleucel | Breyanzi |
| Lomustine | Gleostine |
| Melphalan | Alkeran, Evomela |
| Mercaptopurine | Purixan |
| Mesna | Mesnex |
| Methotrexate | Otrexup; Rasuvo; RediTrex; Trexall; Xatmep |
| Mitomycin | Mutamycin |
| Mitotane | Lysodren |
| Mycophenolate Mofetil | CellCept; Myfortic |
| Natalizumab | Tysabri |
| Nelarabine | Atriance |
| Nivolumab | Opdivo |
| Obinutuzumab | Gazyva |
| Ofatumumab | Arzerra, Kesimpta |
| Osimertinib | Tagrisso |
| Oxaliplatin | ACT Oxaliplatin; Eloxatin; PMS-Oxaliplatin; TARO-Oxaliplatin |
| Paclitaxel | APO-Paclitaxel |
| Palifermin | Kepivance |
| Palivizumab | Synagis |
| Panitumumab | Vectibix |
| Pegademase Bovine | Adagen |
| Pegaspargase | Oncasper |
| Pemetrexed | Alimta; Pemfexy |
| Pentostatin | Nipent |
| Pertuzumab | Perjeta |
| Pimecrolimus | Elidel |
| Plicamycin | Mithracin |
| Pomalidomide | Pomalyst |
| Ponatinib | Iclusig |
| Pralatrexate | Folotyn |
| Prednisone | Multiple |
| Rituximab | Riabni; Rituxan; Ruxience; Truxima |
| Romidepsin | Istodax |
| Sirolimus | Rapamune |
| Streptozocin | Zanosar |
| Sulfasalazine | Azulfidine |
| Talquetemab | Tecartus |
| Trabectedin | Yondelis |
| Tacrolimus | Astagraf XL; Envarsus PA; Prograf; SANDOZ Tacrolimus |
| Temozolomide | Temodar |
| Teniposide | Vumon |
| Thalidomide | Thalomid |
| Thioguanine | Tabloid |
| Thiotepa | Tepadina |
| Tisagenlecleucel | Kymriah |
| Tocilizumab | Actemra |
| Trastuzumab | Herceptin; Herzuma; Kanjinti; Ogiviri; Trazimera |
| Tretinoin | Vesanoid |
| Ustekinumab | Stelara |
| Vincristine | Oncovin; Others |
| Vedolizumab | Entyvio |
| Vorinostat | Zolinza |
| Abatacept | Orenica |
